# Intrinsic capacity predicts negative health outcomes in older adults

**DOI:** 10.1101/2021.05.26.21257829

**Authors:** Erwin Stolz, Hannes Mayerl, Wolfgang Freidl, Regina Roller-Wirnsberger, Thomas M. Gill

## Abstract

**BACKGROUND:** Monitoring trajectories of intrinsic capacity (IC) in older adults has been suggested by the WHO as a means to inform prevention to avoid or delay negative health outcomes. Due to a lack of longitudinal studies, it is currently unclear how IC changes over time and whether repeatedly measured IC predicts negative health outcomes.

**METHODS:** Based on 4,751 repeated observations of IC (range=0-100) during 21 years of follow-up among 754 older adults 70+ years, we assessed longitudinal trajectories of IC, and whether time-varying IC predicted the risk of chronic ADL disability, long-term nursing home stay, and mortality using joint models for longitudinal and time-to-event data.

**RESULTS:** Average IC declined progressively from 77 to 11 points during follow-up, with substantial heterogeneity between older adults. Adjusted for socio-demographics and chronic diseases, a one-point lower IC value was associated with a 7% increase in the risk of ADL disability, a 6% increase in the risk of a nursing home stay, and a 5% increase in mortality. Accuracy for 5- and 10-year predictions based on up to three repeated measurements of IC ranged between moderate and good (AUC = 0.76-0.82).

**DISCUSSION:** Our study indicates that IC declines progressively and that it predicts negative health outcomes among older adults. Therefore, regular monitoring of IC could work as an early warning system informing preventive efforts.

## 1. Introduction

In a shift from a disease-oriented to a functioning- and life-course-oriented approach of healthy aging, the World Health Organization (WHO)[1] introduced the construct of intrinsic capacity (IC), which is defined as a composite measure of a person’s physical and mental capacities. IC is conceptualized as a dynamic construct[2] comprised by five domains (vitality, locomotor, sensory, cognitive, and psychological)[3]. Based on fine-grained measurements and regular monitoring, it may be possible to detect declines in IC ahead of clinical manifestations. This could facilitate the development and testing of preventive interventions to avoid, attenuate or delay the onset of geriatric syndromes and subsequent health care utilization.

Although there is solid evidence[4–7] that indicators associated with IC-domains such as grip strength (vitality) or gait speed (locomotor) predict adverse health outcomes, few studies, to date[8–10], have assessed the predictive ability of the overall IC construct. Moreover, these studies were either cross-sectional[8], or assessed only the association of between-person differences in IC at baseline with the incidence of disability in activities of daily living (ADL)[9,10] and mortality[10]. Thus, it is currently unclear how IC changes over time and whether longitudinal monitoring of IC is informative with regard to the prediction of negative health outcomes. This knowledge is required if IC is to act as an early warning system informing preventive efforts. The aim of this study is therefore to measure changes in IC longitudinally, and to assess whether repeatedly measured IC predicts three negative health outcomes (ADL disability, nursing home stay, and mortality) among community-dwelling older adults.

## 2. Methods

### Data

In the Yale PEP Study[11], monthly telephone interviews and comprehensive home-based face-to-face assessments at 18-month intervals have been conducted since 1998 among 754 community-dwelling health plan members from greater New Haven, Connecticut (US). Eligible participants were aged 70 years or older, and without ADL disability at baseline. For the current study, we used follow-up data through June 2019 (21 years).

### Variables

#### Intrinsic capacity

IC was monitored based on the 18-months assessments, which amounted to a total of 4,751 repeated observations (6.5 observations of IC per person on average). IC was measured via its five domains. (1) *Vitality* was operationalized via muscle strength and respiratory functioning. Muscle strength was assessed by mean hand grip strength (kg) over three readings with a hand-held Chatillon 100 dynamometer. Respiratory functioning was assessed with the maximum peak expiratory flow value (liter/min) over three attempts measured with a Mini-Wright meter. (2) *Locomotor* was assessed with gait speed (in seconds) tested across a 20-foot walk with a turn, the time required to perform three stands in the chair-rising test (in seconds), and the balance test (tandem, semi-tandem, side-by-side, 0-4 points) from the short physical performance battery. (3) The *sensory* domain included near-vision acuity, which was measured with a Jaeger chart (impairment in %), and hearing impairment, which was measured with an Audioscope (impairment in %). (4) *Cognition* was measured with the Mini-Mental-State-Examination (MMSE, range=0-30), and (5) the *psychological* domain was measured with the Center for Epidemiologic Studies Depression scale (CES-D, range=0-11). Input variables were rescaled after stratification by sex with the percent of maximum possible method[12], so that values were comparable longitudinally[13], and ranged from 0 (minimum possible) to 100 (maximum possible). We computed IC in each wave by first calculating a meanscore for domains with multiple indicators (vitality, locomotor) before calculating a meanscore over all five domains. Finally, the obtained meanscore values of IC were again re-scaled so that they ranged from 0-100.

Internal consistency reliability and construct validity of IC was assessed with confirmatory factor analysis (CFA), where we tested a second-order and a bi-factor model[9], with the latter providing the better model fit. We removed hearing impairment due to low correlation (r<.10) to all other IC input variables which led to problematic negative variances in the CFA. Without hearing impairment, maximum-likelihood estimation with robust standard errors via R-package *lavaan* yielded a very good fit for the bi-factor model: e.g. at baseline *χ*^2^(16)=23.56, RMSEA=0.03, CFI=0.99, TLI=0.99, SRMR=0.02. Reliability of the IC as a general factor ranged between acceptable (e.g. at baseline: hierarchical *ω*=0.71) to good (e.g. at 7.5 years: hierarchical *ω*=0.86) during follow-up. The correlation between the extracted IC factor scores and the IC meanscores which were used in the further analysis was high (r=0.93-0.96) throughout follow-up.

#### Negative health outcomes

Negative health outcomes included (1) onset of chronic ADL disability[14] (need for personal assistance in either dressing, bathing, walking inside the house, or transferring from a chair) lasting at least three months, (2) long-term (3+ months) nursing home stay (NHS), and (3) mortality (MOR). ADL and NHS were assessed via monthly telephone interviews, MOR was determined from local obituaries and informants (100% complete). For each outcome, a time-line was created, with the first interview marking the beginning of the observation period until either the time of the outcome, the end of follow-up, or dropout for other reasons, whatever came first.

#### Additional predictor variables

These included sex (men/women), 12 or more years of education (no/yes), age at baseline (years), ethnicity (white/non-white) and the number of chronic diseases at baseline (range=0-9).

### Statistical analysis

To predict the negative health outcomes, we used joint models for longitudinal and time-to-event data[15], an advanced statistical approach where effects of time-varying predictors on time-to-event outcomes are estimated based on their longitudinal characteristics captured with mixed-effects regression models. In contrast to the extended Cox-models for time-dependent predictors, joint models are better suited to handle both incomplete follow-up data and noisy information in time-varying covariates. In the mixed regression sub-model, we first compared different specifications of time according to goodness-of-fit measures (AIC, BIC, LRT-Test) to correctly characterize population- and individual-level IC trajectories[16]. The best fit was obtained with a restricted natural cubic spline with one internal knot placed at the median of the follow-up time capturing non-linear population- and individual-level trajectories of IC. The survival regression sub-model was formulated as:

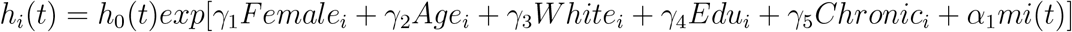

where the risk of an adverse health outcome *h* for individual *i* at time *t* depends on the baseline hazard function *h*_0_ estimated with a cubic-spline (five knots), the regression coefficients *γ*_1−5_ of the time-constant predictors, and the association parameter *α*. For *α*, we used the current value parametrization, i.e. the ‘true’ value of IC based on the individual-level growth model at *t* as a time-varying predictor. To assess the predictive performance, we calculated time-dependent area under the receiver operating characteristic curve (AUC) values using information on IC up to the third year and medium (5 years) and long-term (10 years) prediction windows. AUC values were corrected for overfitting with an internal validation procedure using bootstrap[17]. Joint models were fitted using R-package JMbayes (v0.8-85), and all calculations were done in R: A language and environment for statistical computing (v4.0.3).

## Results

At baseline, average age was 78.4 (SD=5.3, range=70-96) years; 67.1% of the sample were women, 89.5% identified as white, 68.3% had 12 or more years of education, and the median number of chronic diseases reported was 2 (IQR=1). The distribution of IC values at baseline was skewed to the left, i.e. most older adults had initially high levels of IC (Plot A in Figure 1). The mean IC at baseline was 77 (SD=11), which declined progressively to 11 points at the end of follow-up (Plot B in Figure 1). Figure 1 also shows that there was substantial variance between participants (thin grey lines), which increased during follow-up: e.g. IQR at baseline=14, after 6 years=21, and after 12 years=23. Participants who developed chronic ADL disability, who were admitted to a nursing home, or who died during follow-up exhibited faster deterioration in IC on average, relative to those who did not develop the specific outcome (Plot A in Figure 2). Older adults with below-average IC at baseline also had a higher risk of negative health outcomes compared to those with above-average baseline-IC (Plot B in Figure 2).

**Figure 1:**
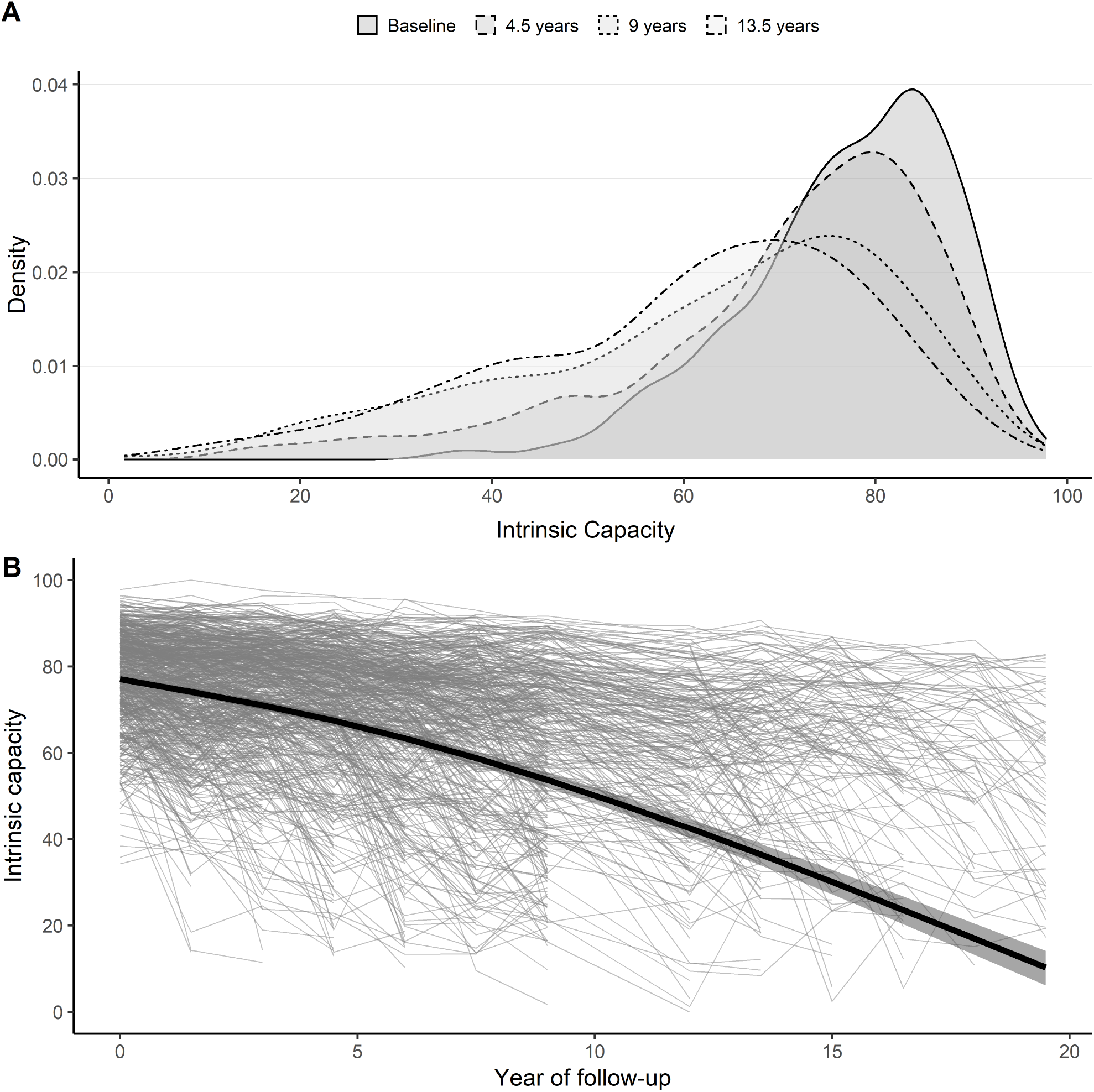
Distribution and trajectory of intrinsic capacity over time. Plot A: Smoothed histograms of the distribution of values of intrinsic capacity at four selected points in time: baseline, 4.5, 9 and 13.5 years. Plot B: Thin grey lines show raw longitudinal observations of intrinsic capacity for 754 participants, which illustrate the broad variety of IC-leves at baseline and of IC-trajectories throughout follow-up (no measurement was available at 10.5 years of follow-up due to lack of funding). Thick black line shows estimated average trajectory of intrinsic capacity from mixed regression sub-model. Shaded areas indicate 95% confidence intervals.

**Figure 2:**
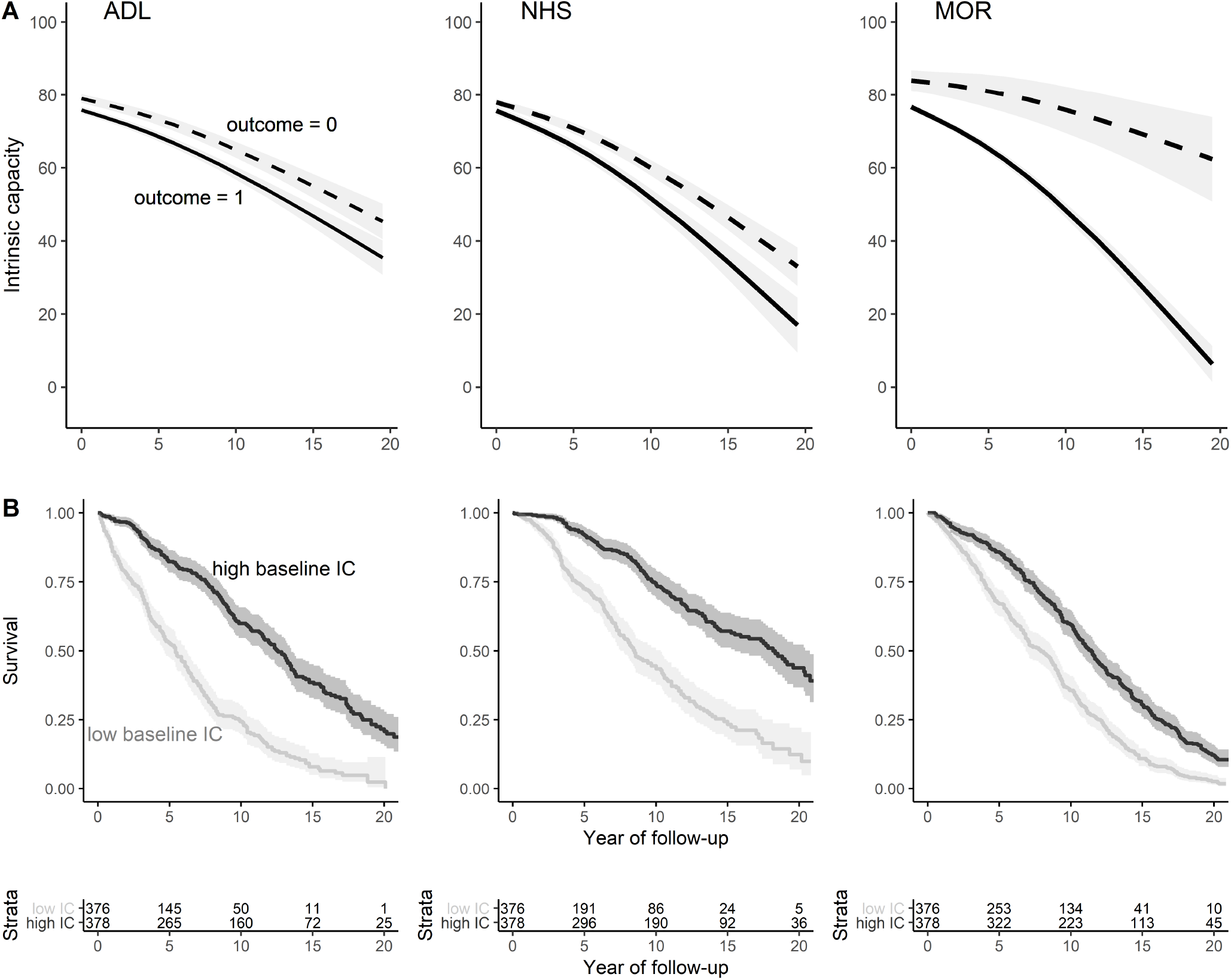
Intrinsic capacity and negative health outcomes. Plot A shows fitted average trajectories from mixed regression models with an interaction term between time and negative health outcome status: solid lines refer to participants who experienced the outcome, dashed lines to participants without the outcome. Only observations of IC before the onset of the outcome were used. Plot B shows Kaplan-Meier plots for survival probability (i.e. the probability to not experience the respective health outcome) by IC category (high = above median IC, low = below median IC) at baseline. Shaded areas indicate 95% confidence intervals. Below plot B, the absolute number of participants at risk by group is shown. ADL = incidence of disability in activities of daily living; NHS = nursing home stay, MOR = mortality, IC = intrinsic capacity.

During follow-up, 61.4% of the participants reported the onset of chronic ADL disability after a median of 5.8 years (IQR=8.1), 42.7% were admitted to a nursing home after a median of 7.3 (IQR=8.8) years, and 93.1% eventually died after a median of 9.4 (IQR=8.6) years. The results from the joint models, adjusted for socio-demographics and chronic diseases, (Table 1) indicate that a one-point lower IC (on scale 0-100) was associated with an 7% (=1/0.94) increase in the risk for ADL (95% CI: 1.06-1.07), a 6% increase in the risk for NHS (95% CI: 1.05-1.07), and a 5% increase in the risk of death (95% CI: 1.04-1.05). The AUC-values indicate that IC, together with information on socio-demographics and chronic diseases, has good discriminative power predicting future negative health outcomes (5-year AUC: ADL=0.81, NHS=0.80, MOR=0.76; 10-year AUC: ADL=0.82, NHS=0.81, MOR=0.76). Figure 3 shows how individual predictions are updated as additional values of IC become available. To illustrate, we selected three female participants aged 74, 81, and 77 years at baseline, of whom B and C developed chronic ADL disability after 11 and 12 years of follow-up, respectively. Their baseline IC values and their IC-trajectories varied considerably, as did their predicted outcome probabilities at different time points. The sharp decline of IC in participant C over time, for example, results in an ever more certain and lower probability to remain free from chronic ADL disability.

**Table 1:** Results from joint models

| Negative outcomes | ADL<br>HR (95% CI) | NHS<br>HR (95% CI) | MOR<br>HR (95% CI) |
| --- | --- | --- | --- |
| Female ($\gamma_1$) | 1.00 (0.83-1.21) | 0.80 (0.63-1.01) | 0.52 (0.45-1.04) |
| Age ($\gamma_2$) | 1.04 (1.02-1.06) | 1.03 (1.02-1.05) | 1.03 (1.01-1.04) |
| White ethnicity ($\gamma_3$) | 1.35 (0.96-1.94) | 1.56 (1.06-2.39) | 1.29 (1.00-1.65) |
| 12+ years of education ($\gamma_4$) | 0.97 (0.80-1.21) | 1.05 (0.84-1.31) | 1.37 (1.16-1.59) |
| Chronic diseases ($\gamma_5$) | 1.06 (0.98-1.15) | 1.14 (1.04-1.25) | 1.16 (1.10-1.24) |
| IC current value ($\alpha$) | 0.94 (0.93-0.95) | 0.94 (0.93-0.95) | 0.95 (0.95-0.96) |
| MODEL INFORMATION |  |  |  |
| Number of participants | 754 | 754 | 754 |
| Number of observations of IC | 3872 | 4507 | 5049 |
| Number of outcomes | 455 (60.3%) | 530 (70.5%) | 702 (93.1%) |
| 5-year AUC | 0.810 | 0.808 | 0.761 |
| 10-year AUC | 0.818 | 0.811 | 0.757 |

Longitudinal effects of joint model not shown. ADL = disability with regard to basic activities of daily living, NHS = nursing home stays, MOR = mortality. $\gamma$ = effect of time-constant predictors, $\alpha$ = effect of time-varying IC, HR = hazard ratio, 95% CI = 95% credible intervals, AUC = time-dependent area under the receiver operating characteristic curve based on information up to year 3 when a majority of participants were still at risk (527 (70%) for ADL, 598 (79%) for NHS, and 653 (87%) for MOR). Number of observations of IC vary across joint models as only observations before the onset of the outcomes were used. Model fitted with MCMC estimation procedure with 3 chains, 50000 iterations per chain, 3000 iterations burn-in, and thinning by 25.

**Figure 3:**
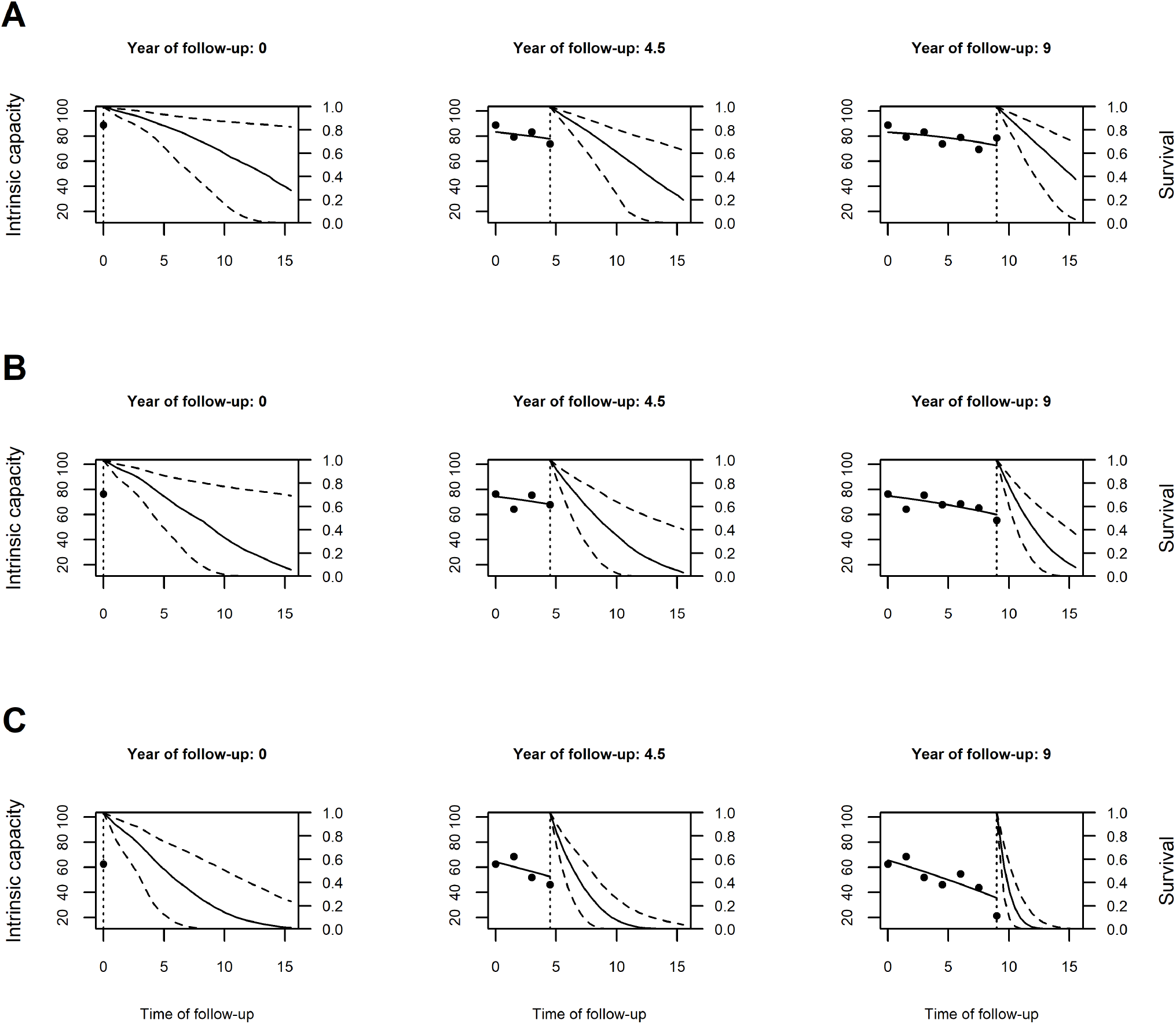
Dynamic predictions of ADL-free status for three selected participants. Trajectories of intrinsic capacity and associated ADL-free survival probabilities for 3 selected participants (A, B, and C) after 1, 4, and 7 repeated IC measurements based on joint model. The x-axis shows time of follow-up in years, the y-axis on the left side represents intrinsic capacity, the y-axis on the right side indicates estimated probability to remain ADL-free. Points are raw observations of IC, the dotted vertical line indicates the last IC-measurement, solid lines left of the dotted vertical line represent IC trajectories (middle and right column), solid lines right of the dotted vertical line refer to trajectories of survival probability to remain ADL-free, and dashed lines show 95% prediction intervals.

## Discussion

IC has been suggested as a fine-grained, comprehensive measure of overall health status to monitor minor health declines in older individuals and populations in order to intervene and subsequently avoid or delay further major health declines. Based on extensive longitudinal data covering more than 20 years among 754 older adults (70+), we showed in this report that IC declined progressively in later life, and that regularly monitoring of IC can predict future negative health outcomes, particularly the onset of chronic ADL disability and NHS, a proxy for severe long-term care needs. These results suggest that monitoring IC could help to target older persons with a heightened risk of care-dependency for preventive interventions.

In the current study, we found that, on average, IC declined progressively over time. However, it is important to keep the substantial between-person heterogeneity around this average trajectory in mind: participants entered the study with broadly varying levels of IC, and IC-trajectories during follow-up varied from stability and even some improvements to drastic declines within a few years. What is more, the between-person variance in IC increased during follow-up. Plans to develop normative trajectories of IC – to identify individual deviations from normality – for preventive actions[2,9] should pay attention to this increasing heterogeneity. In our analysis, we measured IC up to 13 times per person at 18-months intervals. Assessments of IC among older adults during annual health check-ups could provide even more timely information to better tailor diagnostic and therapeutic decisions without additionally taxing routine medical care.

Our study is the first attempt to predict negative health outcomes based on repeatedly measured IC. Whereas previous studies relied on one-time assessments of IC[8–10], we could show that IC declines progressively and the current (or last) IC value predicts future negative health outcomes. The IC therefore could be used to predict probabilities for medium- and long-term negative health outcomes dynamically and to update prognoses, which may act as an early warning system and support medical decision making.

IC has been discussed[2,8] as a concept complementary to or leading up to phenotypic frailty[18]. However, there is a potentially problematic overlap in some of the indicators of frailty and IC (i.e. gait speed, grip strength, items on exhaustion from CES-D), which precluded the inclusion of (pre-)frailty as a negative health outcome in our study. IC also shares a number of characteristics with the other main model of frailty, namely the health deficit accumulation approach and its frailty index (FI)[19]. Both FI and IC are multidimensional – i.e. including information about physical, cognitive and mental health – and continuous indicators of overall health that can be measured repeatedly to assess longitudinal trajectories in the general older population (e.g.[20]). Interestingly, the FI, which is based on 30+ dichotomized health deficits, typically exhibits a right-skewed distribution in community-dwelling samples of older adults, whereas IC values – which are based on fewer but continuous measurements of functioning – exhibited a left-skew in our study. Future studies should compare the predictive performance of the FI and IC for negative health outcomes, considering also the more established conceptual framework and the more straightforward calculation of the FI.

In our analysis, we used joint models for longitudinal and time-to-event data which have two major benefits over extended Cox-regression models. First, the mixed regression sub-model provides unbiased population-level estimates of IC trajectories by making use of all available observations. Second, the joint model approach can handle measurement error in the time-varying predictor. A recent simulation study[16] showed, that when there is a considerable level of measurement error in the time-varying predictor as found in our study by the *ω*-coefficients of IC, joint models provide more accurate estimates compared to the down-biased coefficients from Cox-regression models.

Strengths of this analysis include the long follow-up period and the high number of repeated observations of IC, the use of continuous indicators and assessment of reliability and construct validity of IC, and the statistical approach which accounted for non-linear trajectories of IC, the measurement error in IC, and which yield population-level and individual-level predictions. However, there are also several limitations to this research. First, there is no consensus yet on how to measure IC, with regard both to the selection of indicators[8] and how to calculate, rescale, or validate the summary measure of IC. In this study, we used similar indicators – although no biomarkers were available – and also a bi-factor model as Beard et al.[9] to operationalize IC. However, hearing impairment would not fit into our model, and thus, the domain sensory included only near-vision acuity. Also, there was a considerable level of measurement error in IC, which calls for further clarification of the conceptual model and the measurement strategy. Second, we focused on the overall construct of IC in this study, although our understanding of changes in IC and of potential targets for prevention and treatment could benefit also from assessing the co-evolution of the constituting dimensions of IC over time. Also, different domains likely exhibit different dynamics, e.g. mood likely changes faster or fluctuates compared to more gradual declines in sensory functioning. Third, our data did not include young older adults (50-70 years old), who might be a more appropriate target population for long-term prevention efforts. Fourth, the study participants were members of a single regional health plan 70 years or older who were initially non-disabled. Hence, our results might not be generalizable to the older U.S. population.

In conclusion, our study indicates that IC declines progressively and that regular monitoring of intrinsic capacity can predict negative health outcomes among older adults. IC could therefore work as an early warning system informing preventive efforts. However, more research is needed to improve the conceptual foundations and measurement of IC before routine monitoring can be implemented.

## Data Availability

Requests for access to data from the PEP Study for meritorious analyses from qualified
investigators should be directed to Thomas Gill. The R-Markdown code reproducing all analyses, results and this manuscript are available online.

https://osf.io/h7kyp/

## Conflict of Interest

None declared.

## Data and code availability

Requests for access to data from the PEP Study for meritorious analyses from qualified investigators should be directed to Thomas Gill. The R-Markdown code reproducing all analyses, results and this manuscript are available online (https://osf.io/h7kyp/).

## Funding

This work was supported by a grant from the National Institute on Aging (grant number R01AG17560). Dr. Gill is supported by the Yale Claude D. Pepper Older Americans Independence Center (grant number P30AG21342).

## Acknowledgments

We thank Denise Shepard, Andrea Benjamin, Barbara Foster, and Amy Shelton for assistance with data collection; Geraldine Hawthorne and Evelyne A. Gahbauer for assistance with data entry and data management; Peter Charpentier for design and development of the study database and participant tracking system; and Joanne McGloin for leadership and advice as the Project Director.

## Author Contributions

ES planned the study, performed all statistical analysis, and wrote the article. TMG contributed to the planning of the study, provided access to the data, and critically reviewed the manuscript. HM contributed to the CFA and critically reviewed the manuscript. WF contributed to the planning of the study, and critically reviewed the article. RRW contributed to the interpretation of results and critically reviewed the article.

## Notes

### Competing Interest Statement

The authors have declared no competing interest.

### Author Declarations

The study protocol of the PEP Study was approved by the Yale Human Investigation Committee, and all participants provided verbal informed consent

